# T-SEM of 11 Major Psychiatric Disorders: Identification of Gene Expression Patterns for Cross-Disorder Risk and Drug Repurposing

**DOI:** 10.1101/2022.11.01.22281811

**Authors:** Andrew D. Grotzinger, Kritika Singh, Tyne W. Miller-Fleming, Max Lam, Travis T. Mallard, Yu Chen, Zhaowen Liu, Tian Ge, Jordan W. Smoller

**Affiliations:** Institute for Behavioral Genetics, University of Colorado Boulder, Boulder, CO USA; Department of Psychology and Neuroscience, University of Colorado Boulder, Boulder, CO USA; Division of Genetic Medicine, Department of Medicine, Vanderbilt University Medical Center, Nashville, TN, USA; Vanderbilt Genetics Institute, Vanderbilt University Medical Center, Nashville, TN, USA; Stanley Center for Psychiatric Research, Broad Institute of MIT and Harvard, Cambridge, MA, USA; Analytic and Translational Genetics Unit, Massachusetts General Hospital, Boston, MA, USA; Division of Psychiatry Research, The Zucker Hillside Hospital, Northwell Health, Glen Oaks, NY, USA; Research Division, Institute of Mental Health Singapore, Singapore, Singapore; Human Genetics, Genome Institute of Singapore, Singapore, Singapore; Center for Medical Genetics, School of Life Sciences, Central South University, Changsha, Hunan, China; Psychiatric and Neurodevelopmental Genetics Unit, Massachusetts General Hospital, Boston, MA; Center for Precision Psychiatry, Department of Psychiatry, Massachusetts General Hospital, Boston, MA

## Abstract

**Importance:** Psychiatric disorders display high levels of comorbidity and genetic overlap, necessitating multivariate approaches for parsing convergent and divergent psychiatric risk pathways. Identifying gene expression patterns underlying cross-disorder risk also stands to propel drug discovery and repurposing in the face of rising levels of polypharmacy.

**Objective:** To identify gene expression patterns underlying genetic convergence and divergence across psychiatric disorders along with existing pharmacological interventions that target these genes.

**Design:** This genomic study applied a multivariate transcriptomic method, Transcriptome-wide Structural Equation Modeling (T-SEM), to investigate gene expression patterns associated with four genomic factors indexing shared risk across 11 major psychiatric disorders. Follow-up tests, including overlap with gene sets for other outcomes and phenome-wide association studies, were conducted to better characterize T-SEM results. Public databases describing drug-gene pairs were used to identify drugs that could be repurposed to target genes found to be associated with cross-disorder risk.

**Main Outcomes and Measures:** Gene expression patterns associated with genomic factors or disorder-specific risk and existing drugs that target these genes.

**Results:** In total, T-SEM identified 451 genes whose expression was associated with the genomic factors and 41 genes with disorder-specific effects. We find the most hits for a Thought Disorders factor defined by bipolar disorder and schizophrenia. We identify 39 existing pharmacological interventions that could be repurposed to target gene expression hits for this same factor.

**Conclusions and Relevance:** The findings from this study shed light on patterns of gene expression associated with genetic overlap and uniqueness across psychiatric disorders. Future versions of the multivariate drug repurposing framework outlined here have the potential to identify novel pharmacological interventions for increasingly common, comorbid psychiatric presentations.

---

Psychiatric disorders are highly polygenic and genome-wide analyses have identified hundreds of loci associated with these disorders.^1–3^ However, translating these findings into biological and clinical insights has been difficult. Many causal genetic variants are likely to be expression quantitative trait loci (eQTL) that affect trait variation via modulation of gene expression.^4–6^ Transcriptome-wide association studies (TWAS) can be used to quantity the effect of gene expression itself, the more proximal predictor, on a particular trait. TWAS carries the additional advantage of distilling hundreds of associated genetic variants into the more biologically interpretable subunit of gene-level results. Despite its scientific value, a rate limiting challenge for transcriptomic analyses is the pragmatic difficulty of obtaining gene expression data in relevant tissues, and most such studies of psychiatric disorders have relied on limited resources of postmortem brain tissue samples. Summary-based TWAS circumvents this issue by imputing gene expression for traits for which only genome-wide association study (GWAS) summary statistics are available. This approach has yielded insight into gene expression patterns associated with ADHD,^7^ major depression,^8^ treatment resistant depression,^9^ and autism spectrum disorder.^10^ This is with the interpretive caveat that prior findings do not account for pervasive genetic overlap across disorders.

Many gene expression patterns associated with psychiatric risk are unlikely to show diagnostic specificity given genetic correlations that indicate highly overlapping genetic signal across the psychiatric phenome.^11–13^ Transcriptome-wide Structural Equation Modeling (T-SEM)^14^ is a recently introduced extension of Genomic SEM^15^ for modeling tissue-specific gene expression within a multivariate network of genetically overlapping traits. We apply T-SEM here to investigate the effect of gene expression on four psychiatric genomic factors defined by the shared genetic signal across 11 childhood- and adult-onset psychiatric disorders. We also estimate the T-SEM heterogeneity metric (Q_Gene_), an index of genes with disorder-specific effects that allows for unpacking genetic sources of divergence for correlated traits.

Transcriptomic results can also be used to inform drug repurposing when combined with open-source databases that describe existing drugs that target specific genes. Drug repurposing can circumvent an otherwise costly and time-consuming drug development pipeline^16^ and may propel novel avenues for pharmacotherapy. A transcriptomics informed drug discovery approach has already been applied to identify potential pharmacological interventions for individual psychiatric disorders.^9,17^ Given psychiatric comorbidity that is the norm rather than the exception,^18,19^ it is unsurprising that polypharmacy is equally common, with most help-seeking children^20^ and adults^21^ prescribed ≥ 2 psychiatric medications from a single community visit. As T-SEM identifies gene expression patterns that transcend diagnostic boundaries, these may function as efficacious pharmacological targets that aid in reducing polypharmacy for comorbid or mixed clinical presentations.

## Method

### Univariate TWAS in FUSION

Transcriptomic imputation was performed using the FUSION software^6^ applied to publicly available, European GWAS summary statistics for 11 major psychiatric disorders (Table 1). Imputation was performed using 16 sets of functional weights including the 13 brain-based tissue types from the Genotype-Tissue Expression project (GTEx v8),^22^ the two dorsolateral prefrontal cortex (dlPFC) datasets from the CommonMind Consortium (CMC),^23^ and the prefrontal cortex data from PsychENCODE.^24^ Reference alleles were aligned across univariate GWAS summary statistics and, when this information was available, restricted to SNPs with a minor allele frequency minor allele frequency (MAF) > 1% and an imputation (INFO) score > 0.6. We restricted results to genes with an imputation accuracy (*R*^2^) > 0.7 and only performed imputation for genes with a maximum of 50% of the SNPs with functional weights missing. There were 77,943 tissue-specific gene expression estimates across all 11 disorders.

**Table 1.** *T-SEM Results*

| *Outcome* | Cases | Controls | Population<br>Prevalence | Mean $\chi^2(1)$ | Total Hits | | Unique Genes | |
| --- | --- | --- | --- | --- | --- | --- | --- | --- |
|  |  |  |  |  | Hits | Shared<br>hits | Hits | Shared<br>hits |
| **Factor 1: Compulsive Disorders** | - | - | - | 1.432 | 3 | 3 | 2 | 2 |
| *Factor 1 Q<sub>Gene</sub>* | - | - | - | 1.144 | 2 | 0 | 2 | 0 |
| **Factor 2: Thought Disorders** | - | - | - | 3.317 | 1,085 | 866 | 394 | 315 |
| *Factor 2 Q<sub>Gene</sub>* | - | - | - | 1.530 | 139 | 129 | 31 | 26 |
| **Factor 3: Neurodevelopmental Disorders** | - | - | - | 1.498 | 2 | 2 | 1 | 1 |
| *Factor 3 Q<sub>Gene</sub>* | - | - | - | 1.452 | 4 | 1 | 4 | 1 |
| **Factor 4: Internalizing Disorders** | - | - | - | 1.703 | 132 | 126 | 54 | 53 |
| *Factor 4 Q<sub>Gene</sub>* | - | - | - | 1.167 | 4 | 2 | 4 | 2 |
| Alcohol use Disorder (AUD)<sup>56</sup> | 8,485 | 20,272 | 15.90 | 1.060 | 1 | 0 (1) | 1 | 0 (1) |
| Anorexia Nervosa (AN)<sup>57</sup> | 16,992 | 55,525 | 0.90 | 1.478 | 31 | 3 (0) | 11 | 2 (0) |
| Anxiety Disorders (ANX)<sup>58</sup> | 31,977 | 82,114 | 20.00 | 1.259 | 0 | - | - | - |
| Attention-deficit Hyperactivity Disorder (ADHD)<sup>59</sup> | 19,099 | 34,194 | 5.00 | 1.432 | 17 | 5 (0) | 12 | 5 (0) |
| Autism Spectrum Disorder (ASD)<sup>60</sup> | 18,381 | 27,969 | 1.20 | 1.377 | 3 | 0 (0) | 3 | 0 (1) |
| Bipolar Disorder<sup>61</sup> | 41,917 | 371,549 | 2.00 | 2.246 | 232 | 187 (0) | 106 | 85 (0) |
| Major Depressive Disorder (MDD)<sup>62</sup> | 170,756 | 329,443 | 15.00 | 1.948 | 215 | 163 (0) | 82 | 68 (0) |
| Obsessive Compulsive Disorder<sup>63</sup> | 2,688 | 7,037 | 2.50 | 1.127 | 0 | - | - | - |
| Post-traumatic Stress Disorder (PTSD)<sup>64</sup> | 2,424 | 7,113 | 30.00 | 1.037 | 1 | 0 (1) | 0 | 0 (1) |
| Schizophrenia<sup>65</sup> | 53,386 | 77,258 | 1.00 | 3.427 | 1,306 | 814 (128) | 418 | 291 (24) |
| Tourette's Syndrome<sup>66</sup> | 4,819 | 9,488 | 0.80 | 1.217 | 0 | - | - | - |

*Note.* For the four psychiatric factors and $Q_{\text{Gene}}$ the *Shared Hits* column reports the number of hits that were overlapping with univariate TWAS hits for the individual disorders. Hits for the factors are reported for Bonferroni significant genes that were not significant for $Q_{\text{Gene}}$ . For the 11 disorders, the *Shared Hits* column reports univariate TWAS hits that were overlapping with any of the factor T-SEM hits, along with values in parentheses reporting univariate hits overlapping with the $Q_{\text{Gene}}$ hits. *Hits* and *Shared hits* are reported for: (i) the total cumulative hits across all tissues (*Total Hits*) and (ii) restricting to unique gene IDs when the gene was significant across multiple tissues (*Unique Genes*). Thus, the right-most column of *Unique Genes: Shared Hits* reports shared hits across an individual disorder and psychiatric factor, even if that hit is in two different tissue types. Taken together, Factor 2 had 79 highly unique gene hits (i.e., 394-315) that were not significant in any tissue type for any of the psychiatric disorders. To facilitate comparison across T-SEM and TWAS outcomes, mean $\chi^2$ values reported in each row were converted to $\chi^2(1)$ statistics before taking their means.

### LD-score Regression and Psychiatric Factor Model

GWAS summary statistics were processed with the *munge* function prior to running multivariable LD-score Regression (LDSC)^25^ implemented within Genomic SEM.^15^ Multivariable LDSC produces the genetic covariance matrix and sampling covariance matrix that index genetic overlap and sampling dependencies across estimates, respectively, and are used as input to T-SEM. We used the LD score for the European subsample of 1000 Genomes, excluding the major histocompatibility complex (MHC) region, for LDSC estimation. All heritability estimates were converted to the liability scale by inputting the population prevalence and the sum of the effective sample size.^26^ To produce a set of findings comparable to previous work, we used the same model of these 11 disorders from Grotzinger et al. 2022,^13^ that provided good fit to the current data (CFI = .976, SRMR = .097; Figure 1). This model consists of four correlated genomic factors reflecting subsets of *(i)* Compulsive, *(ii)* Thought, *(iii)* Neurodevelopmental, and *(iv)* Internalizing disorders.

**Figure 1.**
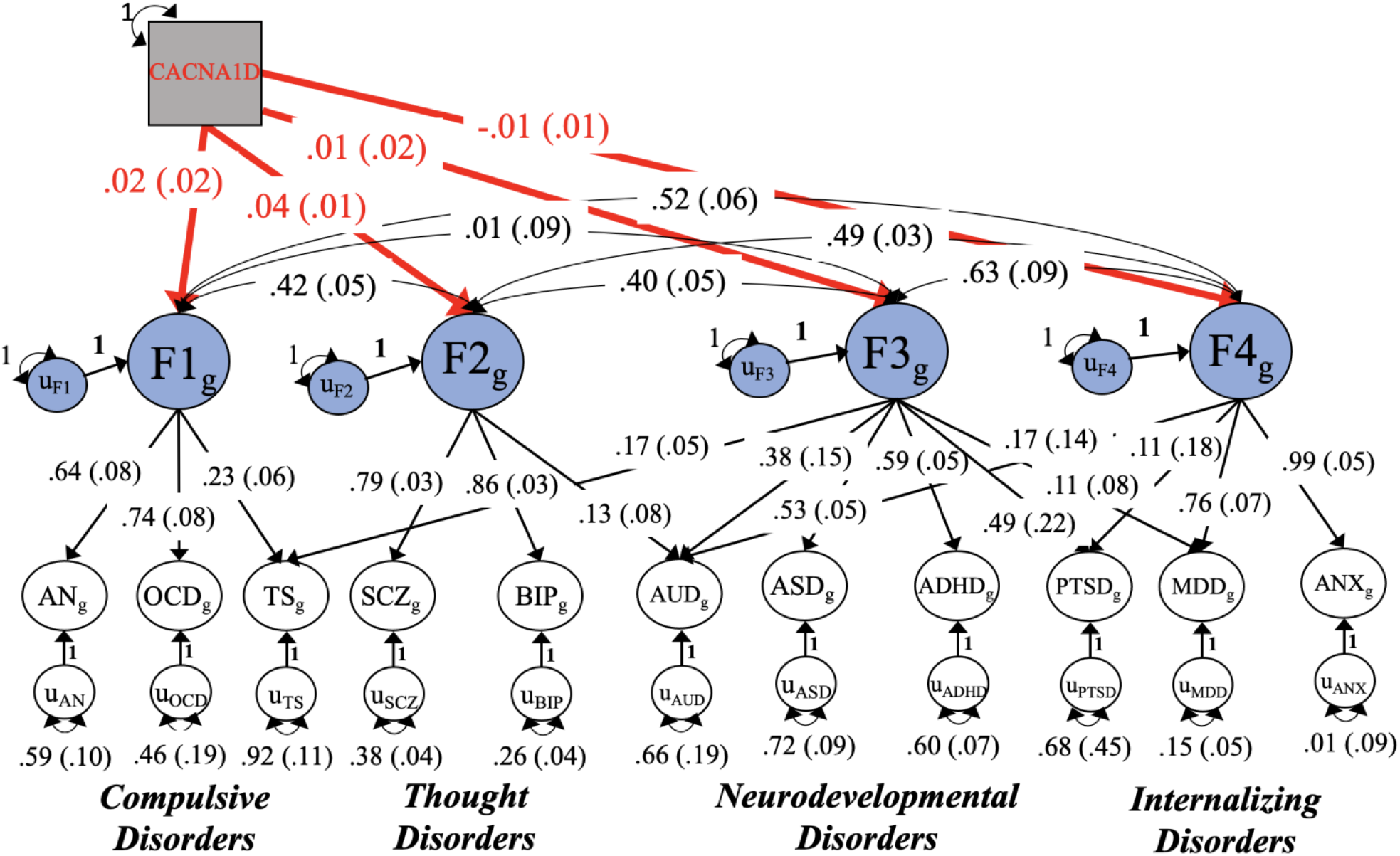
Genomic Factor Model of Psychiatric Disorders. Figure depicts the results from a model in which the gene expression of *CACNA1D* imputed using the PsychENCODE functional weights predicts the four psychiatric factors. CACNA1D was both a top hit for the Thought disorders factor and one of the genes with several identified drug-gene pairs. Depicted results are standardized with respect to the total genetic variance in the 11 psychiatric disorders. The genetic variance of the psychiatric factors reflects 1 + the variance explained by *CACNA1D* expression. Standard errors are shown in parentheses and the effect of gene expression on the factors is highlighted in red. AN = anorexia nervosa; OCD = obsessive-compulsive disorder; TS = Tourette’s syndrome; SCZ = schizophrenia; BIP = bipolar disorder; AUD = alcohol use disorder; ASD = autism spectrum disorder; ADHD = attention-deficit hyperactivity disorder; PTSD = post traumatic stress disorder; MDD = major depressive disorder; ANX = anxiety disorders.

### Transcriptome-wide Structural Equation Modeling

Transcriptome-wide Structural Equation Modeling (T-SEM) is a two-stage approach for examining the effect of tissue-specific gene expression within a multivariate model of genetically overlapping traits. In Stage 1, FUSION and LDSC are applied and the tissue-specific estimates for each gene produced by TWAS are used to expand both the genetic covariance and sampling covariance estimates. The *read_fusion* function was used to scale the univariate TWAS output to be partially standardized gene-phenotype covariances. Genomic inflation was corrected for by multiplying TWAS standard errors by the univariate LDSC intercept when this intercept was > 1. In Stage 2, the effect of tissue-specific gene expression on the four psychiatric factors was estimated. We also produce gene- and factor-specific metrics of heterogeneity via the Q_Gene_ statistic, a metric that identifies genes likely to have disorder specific effects.^14^ Q_Gene_ is estimated as the model χ^2^ difference test comparing a (*i*) a common pathways model in which the gene solely predicts the psychiatric factor to an (*ii*) an independent pathways model in which the gene directly predicts the psychiatric disorders that define the factor.

To better characterize transcriptomic results, we ran five follow-up tests (described in detail in **Online Supplement**). First, Bayesian colocalization analyses were utilized to determine the probability of shared (i.e., colocalized) causal variants across gene expression and the psychiatric factors. Second, the variance explained in nearby multivariate GWAS estimates by gene expression results was calculated. Third, joint analyses were conducted to estimate the conditional significance of physically proximal genes and genes with expression weights across multiple tissues. Conditional findings should be interpreted with caution as genes with nonsignificant conditional associations may simply have higher correlations with other genes. Fourth, Over Representation Analyses (ORA) were conducted using *WebGestalt*^27^ to identify overlap between factor hits and gene sets for external traits. The number of gene expression hits was either too small for these analyses or did not produce significant findings for any outcome except the Thought disorders factor. Fifth, we performed phenome-wide association studies (PheWAS) within the Vanderbilt BioVU European (*n*=70,439) and African Ancestry (*n*=15,174) participants to assess the clinical phenotypes associated with gene expression hits for the psychiatric genomic factors. PheWAS analyses used a strict Bonferroni correction for the number of tested associations across factors, genes, and phecodes (*p* < 1.81E-7). We did not detect any significant associations for individuals of African ancestry, which is likely due to reduced power. Nominally significant associations with psychiatric phenotypes are considered in the **Online Supplement**.

### Multivariate GWAS of Psychiatric Factors

Multivariate GWAS summary statistics were estimated for the psychiatric factors to perform the follow-up colocalization and conditional analyses described above. This was achieved by estimating the SNP effects on the four correlated factors using the *userGWAS* function within Genomic SEM. Prior to running *userGWAS*, the 11 GWAS summary statistics for the disorders were run through the *sumstats* function to align results to the same reference allele, partially standardize estimates with respect to the trait variance, and restrict to MAF > 0.5% and INFO > 0.9 when this information was available. This resulted in a final set of 4,863,931 genetic variants.

### Drug Repurposing

We utilize the Drug-Gene Interaction Database (DGIdb v.4.2.0) and the Broad Institute Connectivity Map (C-MAP) Drug Repurposing Database^28^ to identify existing pharmacological interventions that target T-SEM gene expression hits. These databases include information on the mechanisms of action (MOA) for each drug (e.g., antagonist). This allowed for matching drugs to genes that are likely to have therapeutic, as opposed to adverse, effects based on whether upward or downward expression was associated with psychiatric risk. Oncology, vaccine, and antibody drugs were removed as these forms of drug administration are less likely to be relevant to psychiatric treatment. In line with prior quality control of the DGIdb resource, only drug-gene pairs with interaction scores > 0.5 were retained.^29^ The interaction score reflects the product of supporting publications and the relative drug- and gene-specificity, such that higher values indicate greater support of a drug-gene interaction.^30^ This resulted in a final list of 3,678 and 5,638 drug-gene pairs from DGIdb and C-MAP, respectively. Drug-gene pairs were cross-referenced with the factor and Q_Gene_ hits, though we find no significant results for Q_Gene_. In line with prior drug repurposing pipelines,^31^ we remove genes whose predicted expression was directionally discordant across tissue types for a given factor as the desired MOA is unclear.

## Results

Hits on the factors reflect genes that were significant at *p* < 1.60E-7, reflecting the Bonferroni corrected threshold for the number of tested genes and factors (.05/[77,943_Genes_ × 4_Factors_]), and were not Q_Gene_ significant using the same threshold. As many genes are present across multiple tissues, we focus on results for unique gene IDs.

### Compulsive Disorders Factor

Results revealed 2 hits for the Compulsive disorders factor, both of which were hits for anorexia nervosa (AN): *C3orf62* and *DALRD3* (Supplementary Table 1; Figure 2 for Miami plot). *C3orf62* was also conditionally significant, explained 84.1% of the variance in nearby GWAS results, and was supported by a model of colocalized gene expression and GWAS associations (Supplementary Table 2). There were 2 significant Q_Gene_ hits (*SLC25A27*; *PPAPDC1A*) with directionally opposing effects across AN, obsessive compulsive disorder, and Tourette’s syndrome (Supplementary Table 3; Supplementary Figure 1). No significant PheWAS associations or drug-gene pairs were identified for this factor.

**Figure 2.**
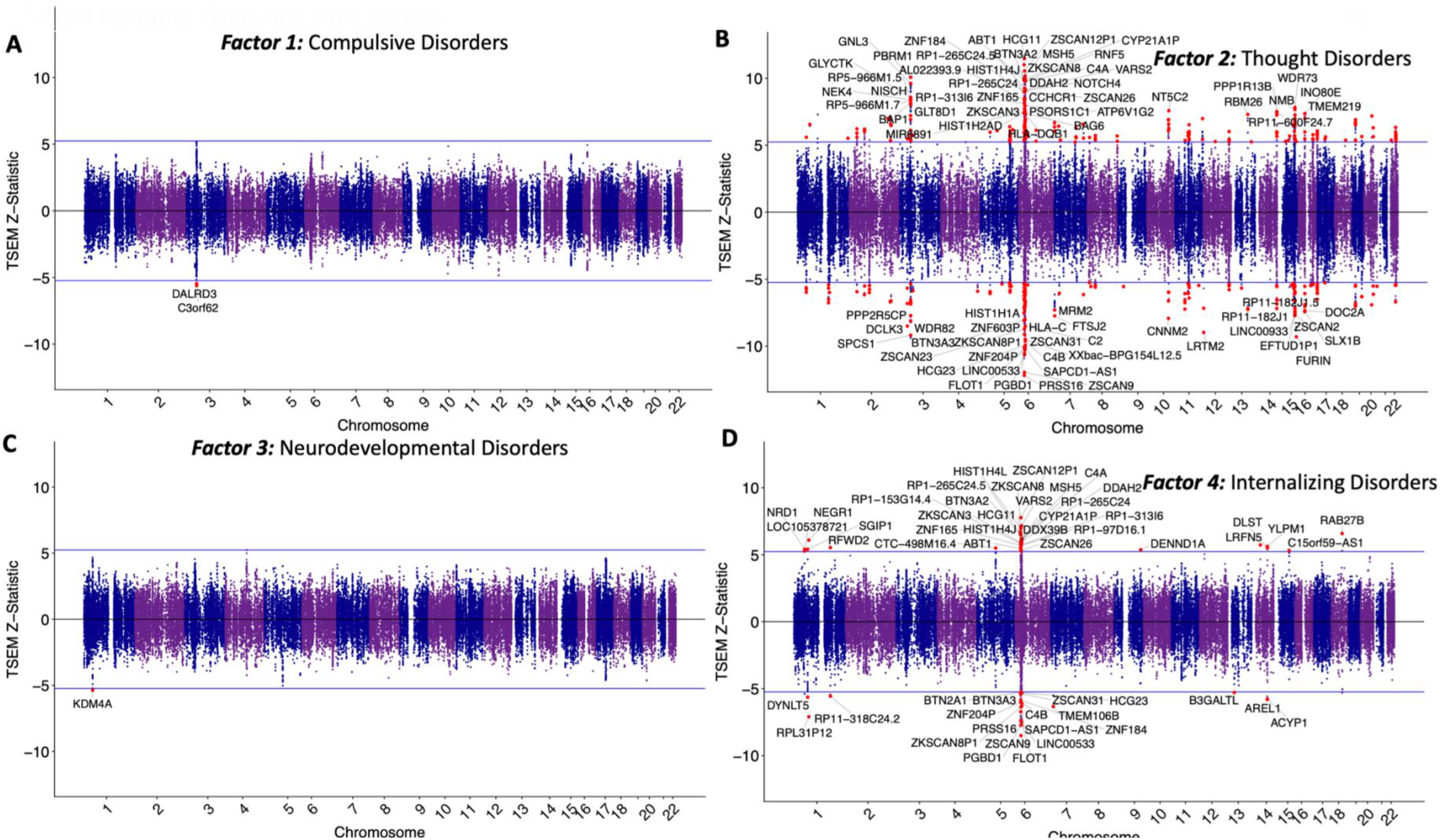
T-SEM of Psychiatric Factors. Panels depict Miami plots of *Z*-statistics for the 77,943 estimated gene expression effects on the Compulsive disorders factor (*Panel A*), Thought disorders factor (*Panel B*), Neurodevelopmental disorders factor (*Panel C*), and Internalizing disorders factor (*Panel D*). *Z*-statistics are signed such that dots on the upper and lower half the plot reflect genes whose upward and downward expression is associated with the psychiatric factors, respectively. The solid blue lines on both sides of the plot reflect the Bonferroni significance threshold, correcting for both the number of tested genes and number of factors. Genes that both surpass these thresholds and are not significant for Q_Gene_ are depicted as red points to denote factor hits. Up to the top 80, Bonferroni significant, unique genes for each factor are labeled.

### Thought Disorders Factor

We identify 394 gene expression hits for the Thought disorders factor defined by bipolar disorder (BIP) and schizophrenia (SCZ; Supplementary Table 4). Highlighting the ability of T-SEM to leverage shared power across genetically overlapping traits, this included 79 novel genes that were not significant for any of the univariate disorder TWAS for any reference tissue. Most hits (63.1%) were supported by a model of colocalized gene expression and GWAS (mean poster probability across all hits = .528), while a smaller percentage (27.3%) indicated independent associations (mean posterior probability = .275). There were 198 conditionally significant factor hits that explained an average of 67.6% of the variance in nearby GWAS estimates (Supplementary Table 5). There were 31 significant Q_Gene_ hits (Supplementary Table 6), 24 of which were significant hits for SCZ.

### WebGestalt

ORA analyses identified significant overlap across Thought Disorders factor hits and eight gene sets, including gene sets for BIP and SCZ (Supplementary Table 7). The most significant gene set was for Alexander Disease, a rare and fatal neurological disorder that causes enlargement of astrocytes.^32^ In addition, there were several autoimmune related gene sets, including: autoimmune thyroid disease, systemic lupus erythematosus, and both immunoglobin A (IgA) deficiency and dysgammaglobulinaemia, which are themselves linked to increased risk for autoimmune diseases.^33^ PheWAS results in BioVU participants of European ancestry revealed 31 significant gene expression-phenotype associations, including 7 genes within the human leukocyte antigen (HLA) region linked to autoimmune diseases such as type I diabetes, multiple sclerosis, and celiac disease (Supplementary Table 8; Supplementary Figure 2). PheWAS results also identified several significant associations between *GAS8* and *CPNE7* expression and several skin cancer phenotypes.

We removed 36 unique genes with a mixture of positive and negative associations across tissue types for the factor hits. This yielded a final list of 358 unique genes associated with the Thought disorders factor for drug discovery follow-up. Drug repurposing results revealed a total of 39 drug-gene pairs across the DGIdb and C-MAP databases (Table 2). These pharmacological interventions may target the shared risk pathways across BIP and SCZ.

**Table 2.** *Drug-Gene Pairs*

| Drug Primary Name | Gene Target | CHR | Mechanism of Action | DGIdb Interaction Score |
| --- | --- | --- | --- | --- |
| RILUZOLE | KCNN3 | 1 | activator | 0.69 |
| AMLODIPINE | CACNA1D | 3 | inhibitor | 0.61 |
| AMLODIPINE MALEATE | CACNA1D | 3 | blocker | 0.61 |
| AZIDOPINE | CACNA1D | 3 | inhibitor | 2.45 |
| DILTIAZEM MALATE | CACNA1D | 3 | blocker | 0.61 |
| ISRADIPINE | CACNA1D | 3 | blocker | 1.4 |
| NIMODIPINE | CACNA1D | 3 | blocker | 1.07 |
| NITRENDIPINE | CACNA1D | 3 | inhibitor | 0.92 |
| CLEVIPREX | CACNA1D | 3 | calcium channel blocker | - |
| DRONEDARONE | CACNA1D | 3 | adrenergic receptor antagonist | - |
| GABAPENTIN | CACNA1D | 3 | calcium channel blocker | - |
| MANIDIPINE | CACNA1D | 3 | calcium channel blocker | - |
| NIFEDIPINE | CACNA1D | 3 | calcium channel blocker | - |
| NILVADIPINE | CACNA1D | 3 | calcium channel blocker | - |
| NISOLDIPINE | CACNA1D | 3 | calcium channel blocker | - |
| SPIRONOLACTONE | CACNA1D | 3 | mineralocorticoid receptor antagonist | - |
| VERAPAMIL | CACNA1D | 3 | calcium channel blocker | - |
| PEMIGATINIB | FGFR1 | 8 | inhibitor | 0.71 |
| NINTEDANIB | FGFR1 | 8 | inhibitor | - |
| PONATINIB | FGFR1 | 8 | inhibitor | - |
| PROGESTERONE | CYP17A1 | 10 | agonist | - |
| LARAZOTIDE ACETATE | CHRNA3 | 15 | partial agonist | 1.59 |
| TC-6499 | CHRNA3 | 15 | agonist | 0.8 |
| VARENICLINE | CHRNA3 | 15 | agonist | 1.36 |
| CYTISINE | CHRNA3 | 15 | acetylcholine receptor agonist | - |
| NICOTINE | CHRNA3 | 15 | acetylcholine receptor agonist | - |
| TETRAMISOLE | CHRNA3 | 15 | immunostimulant | - |
| LORLATINIB | FES | 15 | inhibitor | 1.82 |
| AT-7519 | CDK10 | 16 | inhibitor | 0.61 |
| AZD-5438 | CDK10 | 16 | inhibitor | 0.64 |
| PHA-793887 | CDK10 | 16 | inhibitor | 0.61 |
| RONICICLIB | CDK10 | 16 | inhibitor | 0.64 |
| MK-8353 | MAPK3 | 16 | inhibitor | 1.06 |
| RAVOXERTINIB | MAPK3 | 16 | inhibitor | 3.18 |
| ULIXERTINIB | MAPK3 | 16 | inhibitor | 1.59 |
| TENELIXIMAB | CD40 | 20 | partial agonist | 5.79 |
| BUMETANIDE | SLC12A5 | 20 | inhibitor | - |
| PLUMBAGIN | EP300 | 22 | inhibitor | 0.61 |
| TOSEDOSTAT | XPNPEP3 | 22 | inhibitor | 3.35 |

*Note.* Table reports the 39 drug-gene pairs identified when cross-referencing the DGIdb and C-MAP databases with gene expression hits for the Thought Disorders factor. Results are ordered by chromosome and gene name. Interaction scores are reported for drug-gene pairs from DGIdb. CHR = chromosome.

### Neurodevelopmental Disorders Factor

Results revealed 1 hit for the Neurodevelopmental disorders factor for the *KDM4A* gene that explained 94.4% of the variance in nearby GWAS estimates (Supplementary Tables 9-10). There were 4 hits for Q_Gene_, 3 of which showed moderate, directionally opposing effects across traits (Supplementary Table 11). No significant PheWAS associations or drug-gene pairs were identified for this factor.

### Internalizing Disorders Factor

There were 54 hits for the Internalizing disorders factor, 51 of which overlapped with univariate hits for major depressive disorder (MDD; Supplementary Table 12). Most hits (56.1%) were supported by a model of colocalized gene expression and GWAS (mean posterior probability = .543), while ∼30% were supported by a model of independent expression and GWAS associations (mean posterior probability = .305). There were 25 conditionally significant genes that explained an average of 83.4% of the variance in nearby GWAS estimates (Supplementary Table 13). There were 4 significant hits for Q_Gene_ (Supplementary Table 14). PheWAS results revealed associations with 14 autoimmune phenotypes driven by chromosome 6 genes (Supplementary Table 15). No significant drug-gene pairs were identified.

## Discussion

The current findings reflect a comprehensive transcriptomic examination of 11 major psychiatric disorders and the first strictly multivariate analyses of gene expression patterns associated with psychiatric risk. Applying T-SEM, we identified a cumulative set of 451 genes associated with the genomic factors. Results for the Compulsive and Neurodevelopmental factors were limited, likely reflecting the lower power for the traits defining these factors. Hits for the Internalizing factor included *RPL31P12*, a gene previously shown to be potentially causally associated with MDD^34^ and a top hit for a cross-trait analysis of MDD and insomnia.^35^ Another Internalizing hit, *RAB27B*, was recently associated with MDD, risk for Alzheimer’s disease, and a range of associated outcomes, including increased tangles, β-amyloid and cognitive decline.^36^

Among the top hits for the Thought disorders factor were *NEK4* in the prefrontal cortex, which has been previously linked to BIP and SCZ,^37^ and *ZSCAN2* expression in the prefrontal cortex, which has been associated with SCZ.^38^ Our multivariate findings indicate these prior univariate results likely index gene expression patterns underlying the shared risk pathways across BIP and SCZ. We also observed 24 Q_Gene_ hits that were specific to SCZ, including the *CRHR-IT1* that has been linked to loneliness,^39^ and *VPS29*, a gene uniquely associated with risk for SCZ in prior conditional analyses.^40^ In addition, the *TBC1D5* SCZ and Q_Gene_ hit has been found to interact with *VPS29* to regulate the retromer complex,^41^ a highly conserved subunit associated with early onset Alzheimer’s disease.^42^ These SCZ-specific genes may reflect central components of the divergent risk pathways across SCZ and BIP.

We identified 39 drug-gene pairs for the Thought disorders factor. Lending support for the proposed link between immune dysfunction and psychiatric risk,^43^ several identified drugs were originally developed for autoimmune disorders. For example, Teneliximab was designed as a treatment for autoimmune diseases as a drug that targets the immune system involved *CD40* gene.^44^ In addition, Larazotide Acetate is prescribed for the autoimmune disorder celiac disease.^45^ This subset of drugs is consistent with the ORA and PheWAS follow-up analyses that revealed overlap across Thought Disorders factor hits and autoimmune outcomes. These outcomes included autoimmune thyroid disease, which has itself been linked to increased risk for schizophrenia^46^ and psychosis.^47^

Antihypertensive drugs (e.g., Amlodipine, Nitrendipine, Nimodipine) that target the *CACNA1D* Thought disorders hit were also identified. The *CACNA1D* gene codes for the Ca_v_1.3 α1 subunit, which regulates L-type calcium channels (LTCC) that are increasingly associated with psychiatric risk^48^ and considered viable therapeutic targets.^49^ Consistent with our results that *CACNA1D* is specifically associated with shared genetic risk pathways across BIP and SCZ, LTCC antagonist prescriptions were recently associated with reduced psychiatric hospitalizations for both disorders.^50^ Nimodipine has also been linked to improved cortical efficiency during working memory tasks, indicating this may have therapeutic value for the treatment of negative symptoms, such as cognitive dysfunction.^51^

Finally, we highlight that Riluzole, an antiglutamatergic agent targeting the *KCNN3* hit, was found to be effective in treating negative symptoms in a randomized control trial for schizophrenia.^52^ The therapeutic potential of a pharmacological intervention targeting the glutamatergic system is further supported by increasing evidence linking this system to psychiatric risk.^53^ Prior findings that indicate the potential of Nimodipine and Riluzole to alleviate negative symptoms is exciting as these are typically untreated using current standard approaches.^54^ Within BIP, these two drugs may be especially useful for treating the cognitive deficits more often observed in individuals who experience mania with psychotic features, as compared to those with nonpsychotic mania.^55^

### Limitations and Future Work

Several limitations should be noted. Most importantly, we highlight that results were performed strictly in European samples due to the limited availability of data for other ancestries. It is our hope that in the next few years the expansion of GWAS datasets for different ancestries will reach a sufficient size to extend these findings more representatively. As this work was conducted using common variants with an MAF > 1%, it will also be important to extend this framework and analyses to utilize whole-exome and -genome sequencing datasets. We note also that an analytic pipeline that utilizes a GWAS of the disorders does not account for individual differences in treatment response itself. Future work might then seek to identify the gene expression patterns for individuals with a treatment-resistant form of the disorder. Examining gene expression patterns associated with treatment nonresponse could be used to identify interventions for the subset of individuals most likely to require novel pharmacological approaches.

## Conclusion

We apply T-SEM to 11 major psychiatric disorders to identify patterns of gene expression underlying both genetic convergence and divergence. As GWAS, gene expression, and drug-gene datasets continue to rapidly expand, the multivariate analyses employed here have the potential to identify novel drug targets with relevance for comorbid clinical presentations. Highlighting the promise of T-SEM informed drug repurposing, our analyses identify 39 drugs that target genes associated with the shared risk pathways across BIP and SCZ. Lending support to these findings, a subset of identified drugs have prior evidence from clinical trials indicating their therapeutic potential for one or both disorders. With recent advances in psychiatric genetics, we are finally beginning to deliver on one of the initial promises of psychiatric GWAS to propel drug discovery.

## Data Availability

The data that support the findings of this study are all publicly available or can be requested for access. Specific download links for various datasets are directly below.
Summary statistics for data from the Psychiatric Genomics Consortium (PGC) can be downloaded or requested here:
https://www.med.unc.edu/pgc/download-results/
Summary statistics for the Anxiety phenotype can be downloaded here:
https://drive.google.com/drive/folders/1fguHvz7l2G45sbMI9h_veQun4aXNTy1v
Links to the LD-scores, reference panel data, and the code used to produce the current results can all be found at: https://github.com/GenomicSEM/GenomicSEM/wiki
Links to the functional reference weights from GTEx version 8 and CMC can be found here:
http://gusevlab.org/projects/fusion/
Links to the functional reference weights from PsychENCODE can be found here: http://resource.psychencode.org/
The DGIdb database can be found here (results were downloaded on March 4th, 2022)
https://www.dgidb.org/
The C-MAP Repurposing database can be found her (results were downloaded on March 4th, 2022): https://clue.io/repurposing-app

https://drive.google.com/drive/folders/1fguHvz7l2G45sbMI9h_veQun4aXNTy1v

https://github.com/GenomicSEM/GenomicSEM/wiki

http://gusevlab.org/projects/fusion/

http://resource.psychencode.org/

https://www.dgidb.org/

https://clue.io/repurposing-app

## Code Availability

GenomicSEM software (including the T-SEM extension), is an R package that is available from GitHub at the following URL: https://github.com/GenomicSEM/GenomicSEM

Directions for installing the GenomicSEM R package can be found at: https://github.com/GenomicSEM/GenomicSEM/wiki

The FUSION software used for univariate TWAS analyses is available here: http://gusevlab.org/projects/fusion/

The *WebGestalt* software documentation can be found here and the R package directly installed from the R console: http://www.webgestalt.org/

## Data Availability

The data that support the findings of this study are all publicly available or can be requested for access. Specific download links for various datasets are directly below.

Summary statistics for data from the Psychiatric Genomics Consortium (PGC) can be downloaded or requested here:

https://www.med.unc.edu/pgc/download-results/

Summary statistics for the Anxiety phenotype can be downloaded here: https://drive.google.com/drive/folders/1fguHvz7l2G45sbMI9h_veQun4aXNTy1v

Links to the LD-scores, reference panel data, and the code used to produce the current results can all be found at: https://github.com/GenomicSEM/GenomicSEM/wiki

Links to the functional reference weights from GTEx version 8 and CMC can be found here: http://gusevlab.org/projects/fusion/

Links to the functional reference weights from PsychENCODE can be found here: http://resource.psychencode.org/

The DGIdb database can be found here (results were downloaded on March 4^th^, 2022) https://www.dgidb.org/

The C-MAP Repurposing database can be found her (results were downloaded on March 4^th^, 2022): https://clue.io/repurposing-app

## Acknowledgements

The work presented here was supported by a gift from the Tommy Fuss Fund. ADG was supported by NIH Grants R01MH120219 and RF1AG073593. KS is supported by AHA827137. The dataset(s) used for the BioVU analyses described were obtained from Vanderbilt University Medical Center’s BioVU which is supported by numerous sources: institutional funding, private agencies, and federal grants. These include the NIH funded Shared Instrumentation Grant S10RR025141; and CTSA grants UL1TR002243, UL1TR000445, and UL1RR024975. Genomic data are also supported by investigator-led projects that include U01HG004798, R01NS032830, RC2GM092618, P50GM115305, U01HG006378, U19HL065962, R01HD074711; and additional funding sources listed at https://victr.vumc.org/biovu-funding/. We add that the current analyses would not have been possible without the enormous efforts put forth by the investigators and participants from ENIGMA, the Psychiatric Genetics Consortium, iPSYCH, and UK Biobank.

