## Supplementary material for "T-SEM of 11 Major Psychiatric Disorders: Identification of Gene Expression Patterns for Cross-Disorder Risk and Drug Repurposing": Online Supplement

### T-SEM Follow-up Analyses

*Conditional Analyses.* FUSION offers the ability to conduct follow-up, conditional analyses that aim to tease apart correlated signal across physically proximal genes or the same gene across multiple tissue types. This is achieved by implementing an iterative procedure wherein predictors are added to the FUSION model until there are no remaining significant associations. Conditional analyses were performed using a locus window of 100,000 bp. As $Q_{Gene}$ hits may be physically proximal to factor hits, joint analyses were conducted using the full set of genes that were significant for the factors, including those that were $Q_{Gene}$ significant. This set of findings should be interpreted with caution as conditionally nonsignificant genes may simply be characterized by higher correlations with other genes.

FUSION will also estimate SNP effects conditional on TWAS estimates. These SNP-level conditional analyses produce two primary results: (*i*) the significance of the top SNP in a region after conditioning on TWAS estimates, and (*ii*) variance explained in that region by TWAS estimates. Large portions of SNP-level variance unexplained by TWAS estimates would indicate that the imputed gene expression estimates are reflective of an independent causal feature.

*Colocalization Analyses*. Bayesian colocalization analyses via the *coloc* R package[1] were utilized to determine the probability of shared causal variants across gene expression hits and hits for the corresponding multivariate GWAS of the factor. The resulting output reflects posterior probabilities for five scenarios. Model 0 (PP0) indicates no GWAS or functional association. Model 1 (PP1) examines the probability of only a functional association and Model 2 (PP2) reflects the probability of a GWAS association only. Model 3 (PP3) reflects independent functional and GWAS associations. Model 4 (PP4) is the probability of colocalized functional and GWAS associations. It is important to note that *coloc* assumes that there is a single causal variant, whereas FUSION assumes multiple eQTLs. Consequently, a low posterior probability of Model 3 can also be considered evidence of colocalization, in addition to a high posterior probability for Model 4.

*WebGestalt Analyses.* Over Representation Analysis (ORA) was conducted using *WebGestaltR* (v 0.4.4)[2] in R. Analyses were run using gene sets from eleven human databases, including three gene ontology databases (biological; cellular; molecular processe), three pathway databases (KEGG; Panther; Reactome), two human disease databases (DisGeNet; GLAD4U), two drug databases (DrugBank; GLAD4U) and a single human phenotype database from Human Phenotype Ontology. Analyses were conducted using *WebGestaltR* defaults for ORA analyses, with the exception that the Bonferroni FDR correction for the number of tested gene sets was used to maintain consistency with the multiple testing correction used for T-SEM analyses. *WebGestalt* calculates *p*-values for ORA analyses using a hypergeometric test. The number of gene expression hits was either too small for this analytic pipeline or did not produce significant findings for any outcome except the Thought Disorders factor.

*Clinical Phenotype Association Studies in BioVU.* To examine the clinical implications of the genes identified in our TWAS analysis in an independent population, we calculated genetically regulated gene expression for 85,613 individuals in the Vanderbilt biobank, BioVU. This population consists of individuals who receive care at Vanderbilt University Medical Center and choose to opt-in to the BioVU research study.[3] Genotype information, as well as de-identified electronic medical records, are available for research purposes for individuals, including International Classification of Diseases, ninth and tenth editions (ICD9/10) billing codes. Genotype data for the BioVU population was generated using the Illumina MultiEthnic Genotype Array (MEGAEX). The genotype data were imputed into the HRC reference panel using the Michigan imputation server. Imputed data and the 1000 Genome Project data were combined to carry out principal component analysis (PCA) to identify individuals of European and African ancestry for analysis as previously described.[4] We used the best-performance models from PrediXcan, UTMOST, and JTI approaches trained on GTEx version 8 data to impute the expression of 129 genes within the tissues with the best performance.[5–8] PheWAS was then performed to assess the clinical phenotypes associated with each gene's genetically regulated expression. Phenotypes in BioVU are represented as phecodes, which are assigned as a dichotomous trait and are a hierarchical clustering of the International Classification of Diseases (ICD9/ICD10) codes. For each phenotype, we required a minimum number of 20 cases for inclusion in our PheWAS analyses, which resulted in testing 1,704 phecodes in 70,439 individuals of European ancestry and 1,361 phecodes in 15,174 individuals of African ancestry.

We used the *PheWAS* package in *R* to perform logistic regression to identify the phecodes that are significantly associated with imputed gene expression after adjusting for sex, current age, median age of medical record, genotype batch, and the top ten principal components from genetic data to control for population stratification.[9] For Factor 2 (Thought disorders) and Factor 4 (Internalizing disorders) we only utilize conditionally significant genes in order to reduce the number of tested associations given the large number of hits for these two factors. Analyses were also restricted to unique gene IDs across tissue types resulting in two tested genes for Factor 1, 140 for Factor 2, 1 for Factor 3, and 19 for Factor 4. We employ a Bonferroni for the number of tests performed across all factor (i.e., 0.05/[1,704 phenotypes × 162 genes tested across factors]) to determine statistical significance.

*PheWAS for Psychiatric Phecode.* We examined the top psychiatric phenotype associations for the gene expressions from each factor to assess whether the phenotypes used within the T-SEM were recapitulated in the BioVU participants. The top associations between the Factor 1 (Compulsive disorders) genes, in individuals of European ancestry included nominal associations between decreased expression of *DALRD3* and *C3orf63* with senile dementia ($p$ = 3.71E-4), tobacco use disorder ($p$ = 4.24E-4), and substance addiction and disorders ($p$ = 0.023). Decreased *DALRD3* was also nominally associated with obsessive-compulsive disorders ($p$ = 0.023). In individuals of African ancestry, we also identified nominal associations between decreased *DALRD3* expression and alcohol-related disorders ($p$ = 0.024), in addition to anxiety disorders ($p$ = 0.005) and autism ($p$ = 0.036).

Factor 2 (Thought disorders) phenotype associations in individuals of European ancestry included increased expression of *XPNPEP3* with paranoid disorders ($p$ = 1.36E-5) and decreased expression of *NEK4* with eating disorder ($p$ = 2.73E-4). In individuals of African ancestry, increased expression of *KCNN3* and *FURIN* was nominally associated with schizophrenia ($p$ = 1.889E-4 and $p$ = 0.018, respectively).

Increased expression of *DMRTA1* in individuals of European ancestry (Factor 3: Neurodevelopmental disorders) was nominally associated with senile dementia and mental disorders due to brain damage ($p$ = 0.006). Interestingly, increased expression of *DMRTA1* in individuals of African ancestry was nominally associated with pervasive developmental disorders and ADHD (*p* = 1.494E-3 and *p* = 5.307E-3), while decreased *DMRTA1* expression was nominally associated with PTSD (*p* = 0.011).

Factor 4 (Internalizing disorders) nominal associations within individuals of European ancestry included decreased expression of *LRFN5* with hallucinations (*p* = 4.10E-3) and ADHD (*p* = 4.24E-3) and increased expression of *LRFN5* with acute reaction to stress (*p* = 0.010). Nominal associations of Factor 4 in individuals of African ancestry included decreased expression of *ARSA* with tobacco use disorder (*p* = 0.003) and anxiety disorder (*p* = 0.030), and increased *TMEM106B* expression with PTSD (*p* = 0.033).

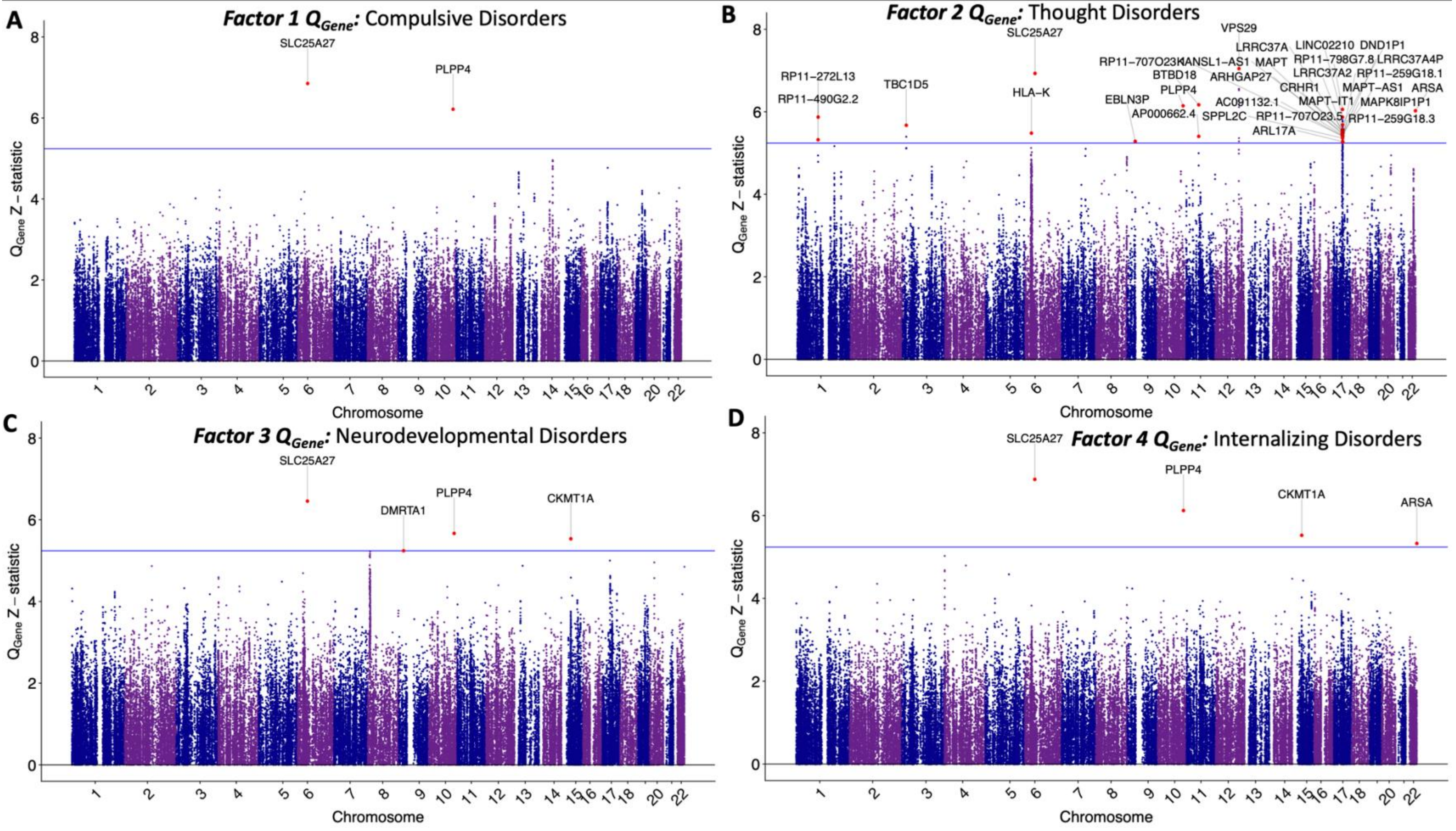


**Supplementary Figure 1. $Q_{Gene}$ Manhattan Plots across Psychiatric Factors.** Panels depict Manhattan plots of factor-specific $Q_{Gene}$ Z-statistics for the 77,943 examined genes for the Compulsive disorders factor (*Panel A*), Thought disorders factor (*Panel B*), Neurodevelopmental disorders factor (*Panel C*), and Internalizing disorders factor (*Panel D*). Z-statistics are not signed here as $Q_{Gene}$ is calculated using model chi-square comparisons that are themselves unsigned. The solid blue line reflects the Bonferroni significance threshold, correcting for both the number of tested genes and number of factors. Genes that surpass this threshold are labeled and depicted as red points.

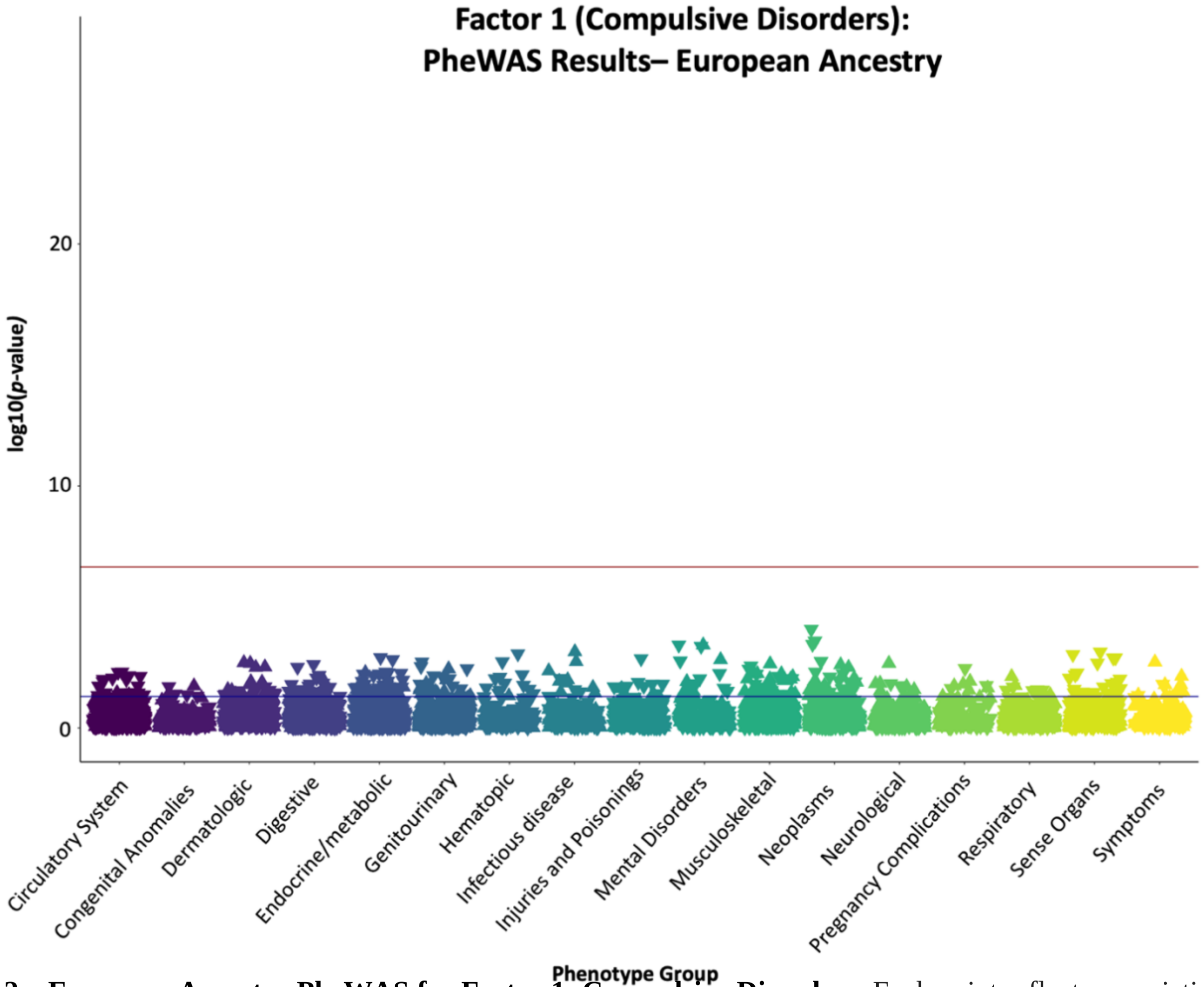


**Supplementary Figure 2a. European Ancestry PheWAS for Factor 1: Compulsive Disorders.** Each point reflects associations between the different phecodes in BioVU and imputed gene expression for genes found to be significantly associated with Factor 1 in the current analyses. Results are grouped and color-coded on the x-axis according to different phenotype groups and vertically positioned on the y-axis according to their -log10(p-values) in the PheWAS. The y-axis is scaled to be consistent across all PheWAS plots for comparative purposes. Upward and downward triangles indicate positive and negative gene-phecode associations, respectively. The black line indicates a threshold of $p < .05$ and the red line the Bonferroni corrected threshold used to define statistical significance in the current analyses.

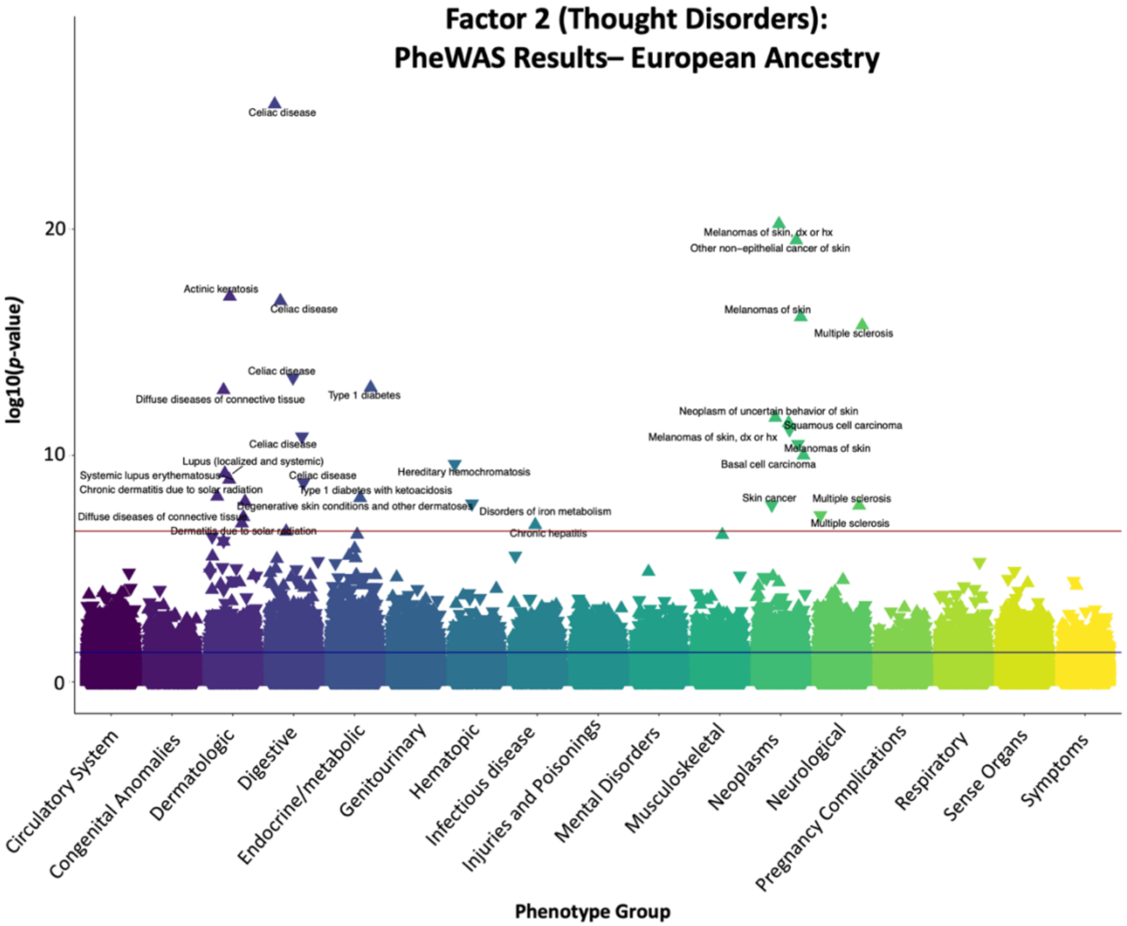


**Supplementary Figure 2b. European Ancestry PheWAS for Factor 2: Thought Disorders.** Each point reflects associations between the different phecodes in BioVU and imputed gene expression for genes found to be significantly associated with Factor 2 in the current analyses. Results are grouped and color-coded on the x-axis according to different phenotype groups and vertically positioned on the y-axis according to their -log10(p-values) in the PheWAS. The y-axis is scaled to be consistent across all PheWAS plots for comparative purposes. Upward and downward triangles indicate positive and negative gene-phecode associations, respectively. The black line indicates a threshold of $p < .05$ and the red line the Bonferroni corrected threshold used to define statistical significance in the current analyses. Statistically significant gene-phecode associations above this threshold are labeled. Duplicate labels (e.g., for celiac disease) indicate separate gene-phecode associations for different Factor 2 gene hits.

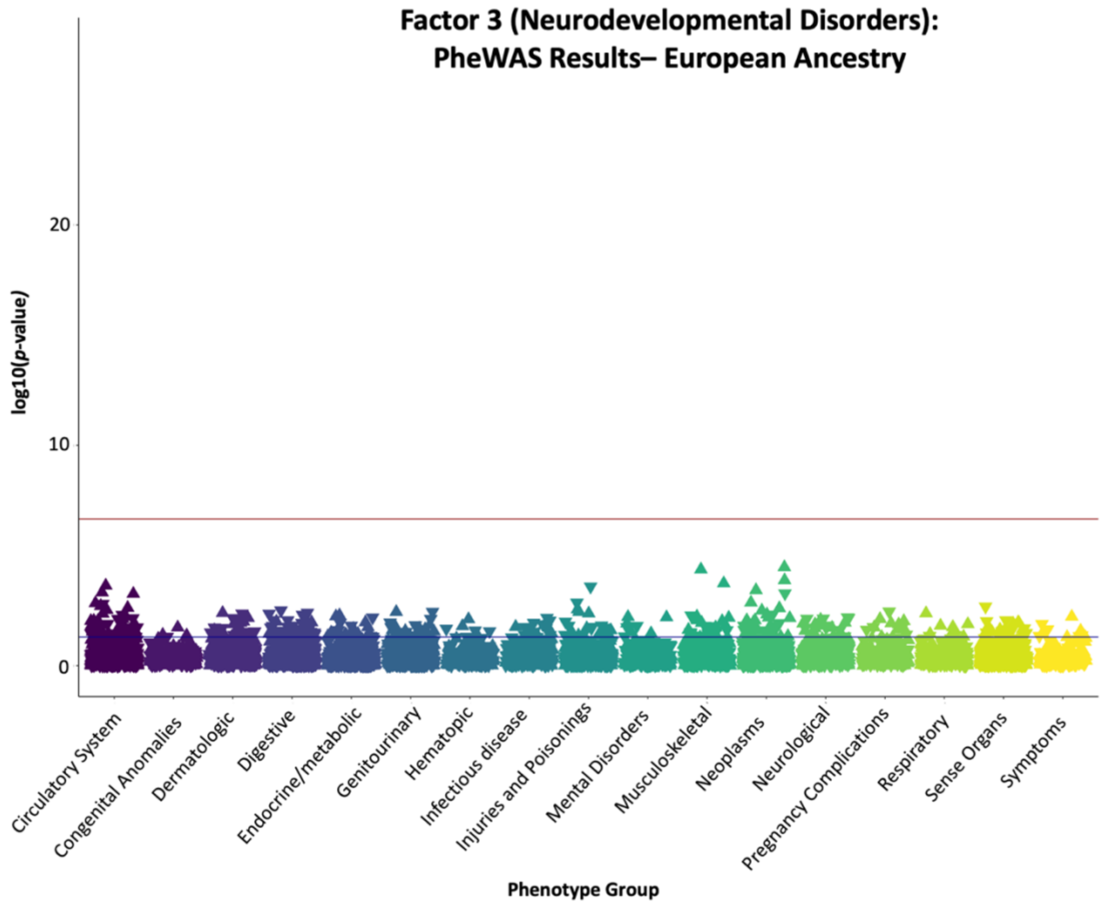


**Supplementary Figure 2c. European Ancestry PheWAS for Factor 3: Neurodevelopmental Disorders.** Each point reflects associations between the different phecodes in BioVU and imputed gene expression for genes found to be significantly associated with Factor 3 in the current analyses. Results are grouped and color-coded on the x-axis according to different phenotype groups and vertically positioned on the y-axis according to their -log10(p-values) in the PheWAS. The y-axis is scaled to be consistent across all PheWAS plots for comparative purposes. Upward and downward triangles indicate positive and negative gene-phecode associations, respectively. The black line indicates a threshold of $p < .05$ and the red line the Bonferroni corrected threshold used to define statistical significance in the current analyses.

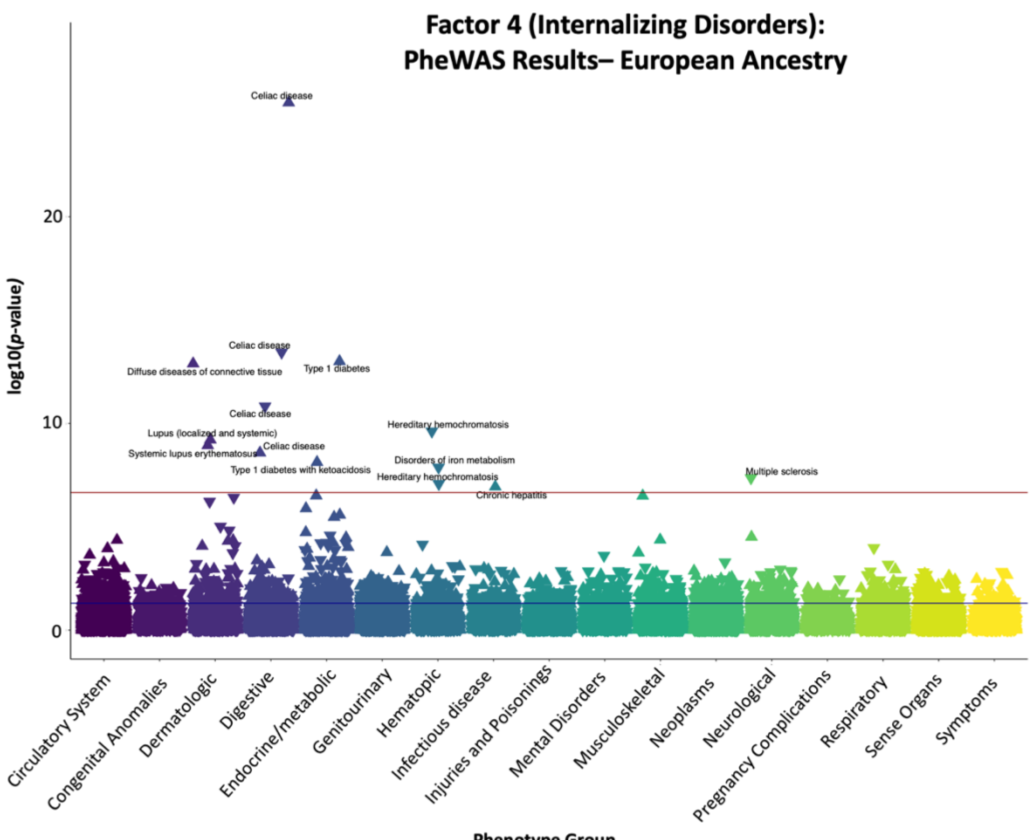


**Supplementary Figure 2d. European Ancestry PheWAS for Factor 4: Internalizing Disorders.** Each point reflects associations between the different phecodes in BioVU and imputed gene expression for genes found to be significantly associated with Factor 4 in the current analyses. Results are grouped and color-coded on the x-axis according to different phenotype groups and vertically positioned on the y-axis according to their -log10(p-values) in the PheWAS. The y-axis is scaled to be consistent across all PheWAS plots for comparative purposes. Upward and downward triangles indicate positive and negative gene-phecode associations, respectively. The black line indicates a threshold of $p < .05$ and the red line the Bonferroni corrected threshold used to define statistical significance in the current analyses. Statistically significant gene-phecode associations above this threshold are labeled. Duplicate labels (e.g., for celiac disease) indicate separate gene-phecode associations for different Factor 4 gene hits.

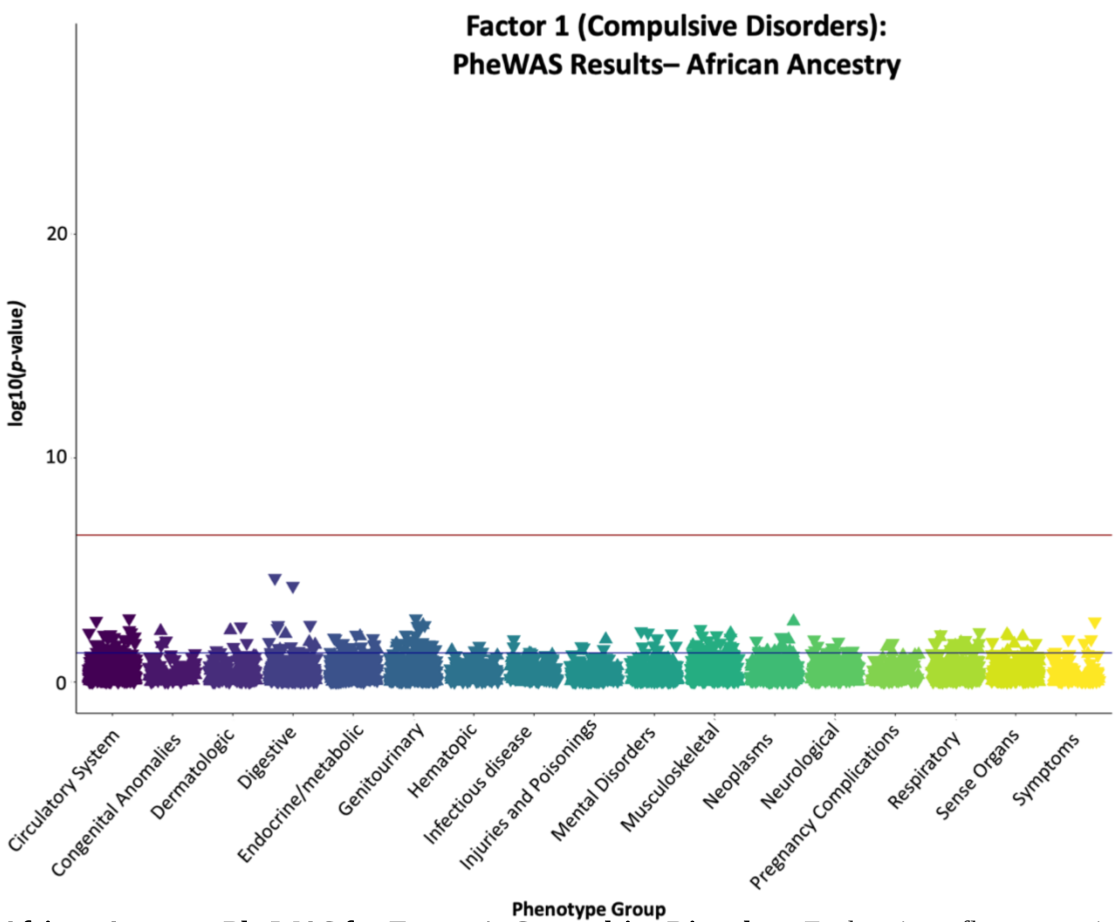


**Supplementary Figure 3a. African Ancestry PheWAS for Factor 1: Compulsive Disorders.** Each point reflects associations between the different phecodes in BioVU and imputed gene expression for genes found to be significantly associated with Factor 1 in the current analyses. Results are grouped and color-coded on the x-axis according to different phenotype groups and vertically positioned on the y-axis according to their -log10(p-values) in the PheWAS. The y-axis is scaled to be consistent across all PheWAS plots for comparative purposes. Upward and downward triangles indicate positive and negative gene-phecode associations, respectively. The black line indicates a threshold of $p < .05$ and the red line the Bonferroni corrected threshold used to define statistical significance in the current analyses.

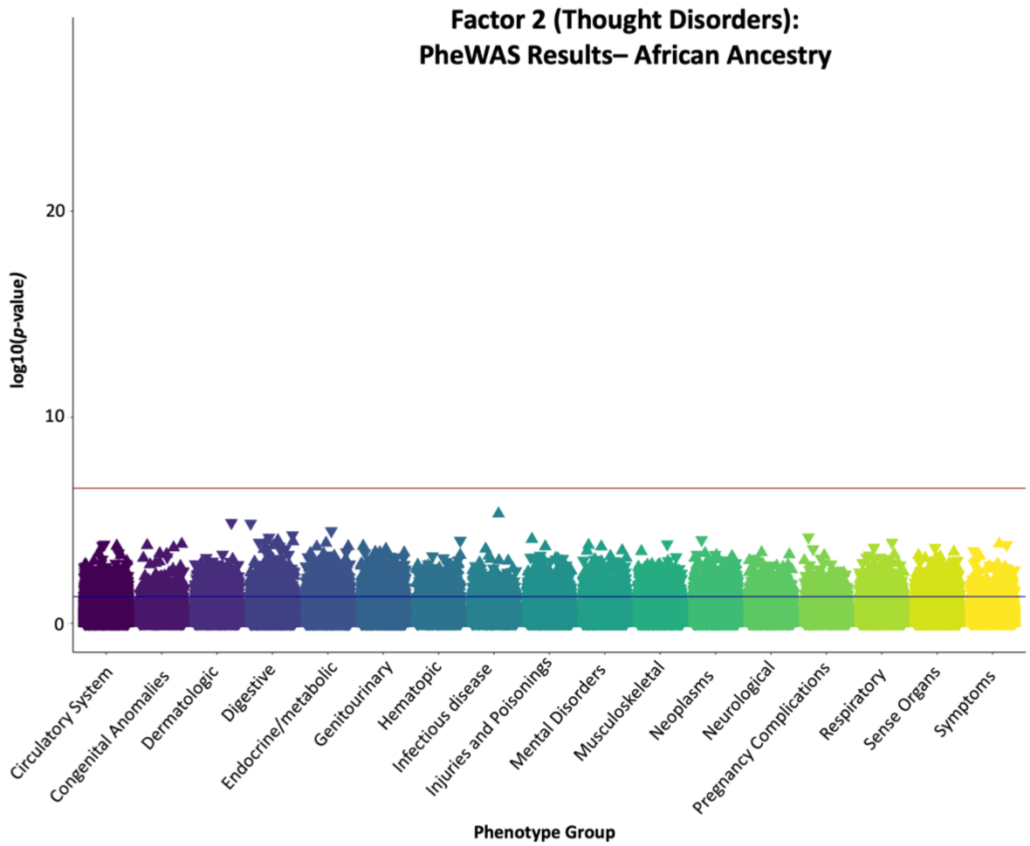


**Supplementary Figure 3b. African Ancestry PheWAS for Factor 2: Thought Disorders.** Each point reflects associations between the different phecodes in BioVU and imputed gene expression for genes found to be significantly associated with Factor 2 in the current analyses. Results are grouped and color-coded on the x-axis according to different phenotype groups and vertically positioned on the y-axis according to their -log10(p-values) in the PheWAS. The y-axis is scaled to be consistent across all PheWAS plots for comparative purposes. Upward and downward triangles indicate positive and negative gene-phecode associations, respectively. The black line indicates a threshold of $p < .05$ and the red line the Bonferroni corrected threshold used to define statistical significance in the current analyses.

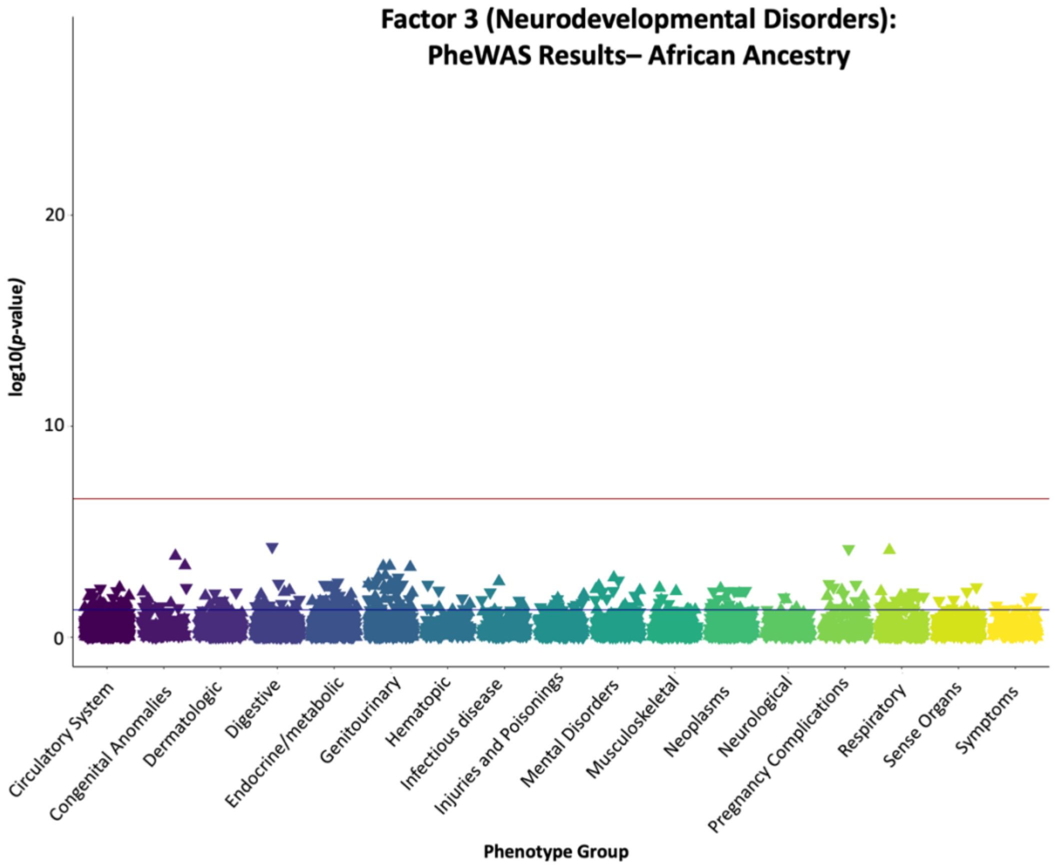


**Supplementary Figure 3c. African Ancestry PheWAS for Factor 3: Neurodevelopmental Disorders.** Each point reflects associations between the different phecodes in BioVU and imputed gene expression for genes found to be significantly associated with Factor 3 in the current analyses. Results are grouped and color-coded on the x-axis according to different phenotype groups and vertically positioned on the y-axis according to their -log10(p-values) in the PheWAS. The y-axis is scaled to be consistent across all PheWAS plots for comparative purposes. Upward and downward triangles indicate positive and negative gene-phecode associations, respectively. The black line indicates a threshold of $p < .05$ and the red line the Bonferroni corrected threshold used to define statistical significance in the current analyses.

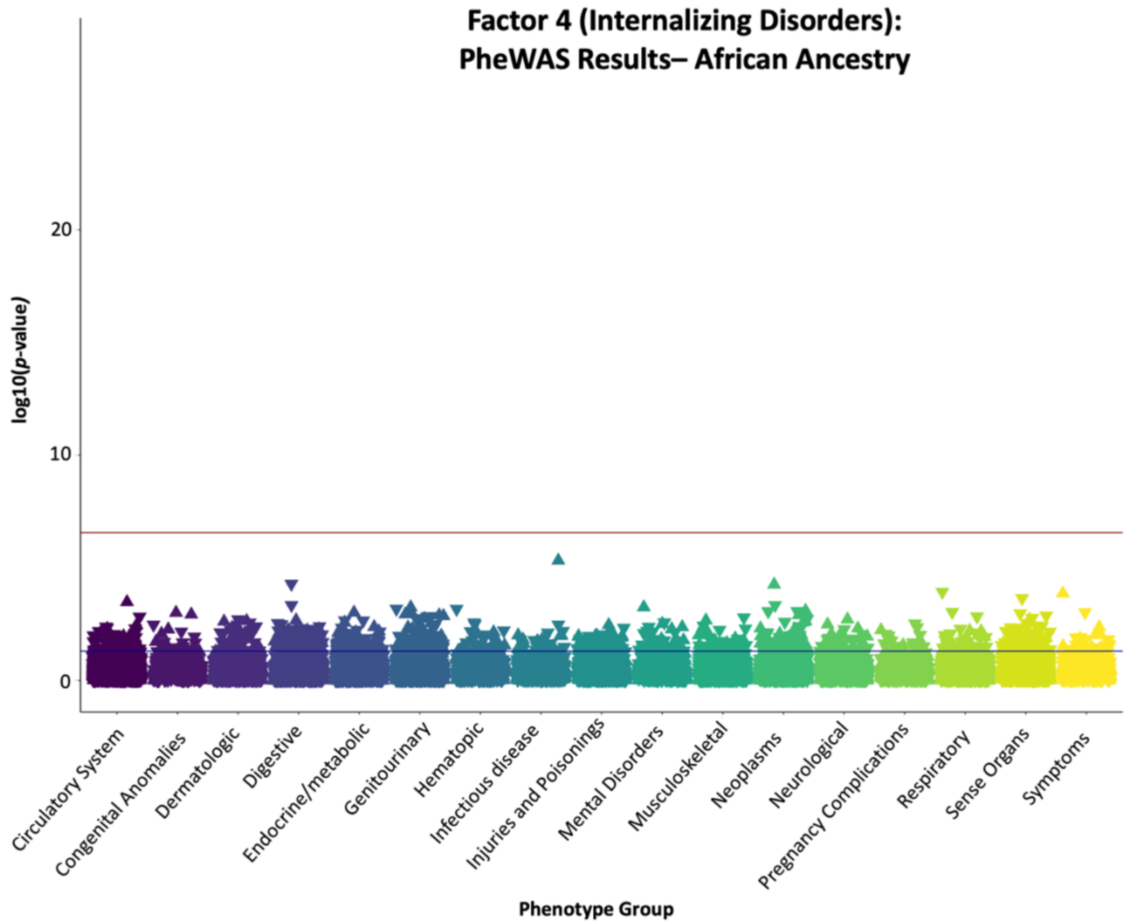


**Supplementary Figure 3d. African Ancestry PheWAS for Factor 4: Internalizing Disorders.** Each point reflects associations between the different phecodes in BioVU and imputed gene expression for genes found to be significantly associated with Factor 4 in the current analyses. Results are grouped and color-coded on the x-axis according to different phenotype groups and vertically positioned on the y-axis according to their -log10(p-values) in the PheWAS. The y-axis is scaled to be consistent across all PheWAS plots for comparative purposes. Upward and downward triangles indicate positive and negative gene-phecode associations, respectively. The black line indicates a threshold of $p < .05$ and the red line the Bonferroni corrected threshold used to define statistical significance in the current analyses.
